# A Personalized Multi-Component Lifestyle Intervention Program Leads to Improved Quality of Life in Persons with Chronic Kidney Disease

**DOI:** 10.1101/19007989

**Authors:** Samuel A. Headley, Jasmin C. Hutchinson, Brian A. Thompson, Marissa L. Ostroff, Courtney J. Doyle-Campbell, Allen E. Cornelius, Kristen Dempsey, Jennifer Siddall, Emily M. Miele, Elizabeth E. Evans, Brianna Wood, Cherilyn M. Sirois, Brett A. Winston, Stefanie K. Whalen, Michael. J. Germain

## Abstract

**Introduction:** Lifestyle interventions have been shown to produce favorable changes in some health outcomes in patients with chronic kidney disease (CKD). However, few such studies, employing “real world” methods have been completed in patients with CKD.

**Objective:** This study tested the effectiveness of a comprehensive, multicomponent, lifestyle intervention, delivered through individualized counseling on a variety of health outcomes in pre-dialysis CKD patients.

**Methods:** Eligible patients were assigned randomly to the intervention (TR) or usual care group (UC). A six-month home-based program involving personalized counseling to increase physical activity to recommended levels among stage G3a to G4 CKD patients while exchanging plant proteins for animal proteins was implemented. Physical function, cardiovascular function, dietary intake, medication use, and health-related quality of life (HRQOL) were assessed at baseline and after 1-month, 3-months (M3) and 6-months (M6).

**Results:** Forty-two, patients (age 60.2 ± 9.2, BMI 34.5 ± 7.8) participated in this study (TR=27 UC=15). The intervention reduced (p<0.05) brachial (bSBP) and central systolic blood pressures (cSBP) at month 3 (M3) but both were attenuated at month 6 (M6). Scores on the effect of kidney disease subscale of the HRQOL measure improved in the intervention group at M3 and M6. There was no change in the other measures of HRQOL or in any physical function scores.

**Conclusions:** This personalized multi-component lifestyle intervention enabled CKD patients to self-report fewer concerns with how CKD affected their daily lives independent of changes in physical function.

## Introduction

Thirty million adults within the United States are estimated to have chronic kidney disease (CKD)(1),(2). Lifestyle interventions can improve health outcomes in chronic diseases(3)(4). Navaneethan et al(2) reported that a 1-year pilot program among predialysis participants led to a 6% reduction in weight that was associated with favorable changes in various biomarkers. They also showed a slower estimated glomerular filtration rate (eGFR) decline in the intervention group(2). Howden et al.(3) examined the impact of a lifestyle intervention on cardiorespiratory fitness (CRF) and risk factors of cardiovascular disease (CVD) in stage 3-4 CKD participants. They found that CRF can be improved in CKD participants when appropriate lifestyle interventions are employed(3).

Among the studies that have incorporated lifestyle interventions involving CKD participants, apart from Howden et al.,(5) most, have focused on diet and exercise interventions only(3)(6). Few have utilized behavioral interventions, although some have also included behavioral counseling, delivered in a group setting Howden et al., 2013, (5) or individually via telephone (7). Few have reported using behavioral counseling delivered in person on an individual basis. Furthermore, many have used in-center programs which have the advantage of offering direct patient supervision, but may not be practical to implement in clinical practice. Providing participants with instructions to follow at home is seen as a pragmatic approach(8).

The aim of this 6-month randomized controlled trial was to determine the effectiveness of an individualized, comprehensive, lifestyle intervention program including dietary, physical activity, pharmaceutical, and behavioral counseling, on physical function, cardiovascular function, dietary intake, medication adherence, self-efficacy, and health-related quality of life (HRQOL) in a sample of pre-dialysis CKD patients. We hypothesized that individuals who participated in the intervention program (TR) would show greater improvements in all dependent variables compared to individuals in the usual care (UC)group.

## Materials & Methods

Participants were recruited from a nephrology practice in Western Massachusetts and were cleared by their nephrologists prior to participation. Eligible (stage G3a-G4, eGFRs 15-59 ml/min/1.73m^2^ 18-75 years, and currently inactive) participants signed a consent form that had been approved by the Institutional Review Boards of both Springfield College and Western New England University. Exclusionary criteria included contraindications to exercise as defined by the American College of Sports Medicine (9), current use of patiromer, contraindication to patiromer use, and current involvement in an exercise program.

Individuals were assigned randomly to either TR or UC in a 2:1 ratio. Serum potassium (K^+^) levels were monitored on a monthly basis since participants were encouraged to adopt a plant-based diet. For individuals in TR, if K^+^ was greater than 5.2 mEq/L, they were prescribed patiromer according to the manufacturer’s recommendations. UC received the current standard of care. They were allowed to take patiromer, if prescribed independently by their physician.

### Testing

Participants were tested on four occasions; baseline (BL), month 1 (M1), month 3 (M3) and month 6 (M6). During the morning of each test day, participants were assessed for anthropometrics, brachial (bBP), central blood pressures (cBP), augmentation index (AIx@75) using the SphygmoCor Excel device (SphygmoCor, AtCor Medical, Sydney, Australia). Other assessments included medication adherence, using the Morisky 8-point scale (MMAS-8), the functional movement screen (FMS), the 6-minute walk test (6MWT), the short physical performance battery (SPPB), and leg extension, leg curl strength, and power using a computerized dynamometer (Biodex Medical Systems, Shirley, NY). Participants also completed the Kidney Disease and Quality of Life (KDQOL) - Short Form (10), the Self-Efficacy to Regulate Exercise Habits Questionnaire (11), and the Self-Efficacy to Regulate Eating Habits Questionnaire. (12) Identical procedures were followed at BL, M1 and M3. At BL and M6, when blood samples were taken for (K^+^), some was also used for the assessment of other biomarkers.

### Blood parameter analysis

The inflammatory biomarkers C-reactive protein (CRP) and interleukin-6 (IL-6) were quantified by immunoassay using commercially available kits. Other analytes (i.e., K^+^ and Phos) were analyzed using standard laboratory methods.

### Individual counseling

Following testing sessions, individuals assigned to TR received a counseling intervention. These participants were instructed as to how to prepare a plant-based meal and subsequently ate a plant-based meal for lunch. Following lunch, participants received one-on-one counseling for nutrition, physical activity, pharmacy, and behavior change. UC were told to follow the normal instructions of their nephrologist.

### Nutrition

Participants kept a 3-day food log prior to each visit. The logs were analyzed and then individually reviewed with the participant during their nutritional counseling session. Specific attention was paid to the amount and quality of protein, sodium (Na) and Phos in the diet. Recommendations for dietary modifications were based on a vegan approach. Participants in TR were encouraged to decrease the animal protein consumed and choose more plant-based meals. Individual barriers to success in achieving the suggested dietary modifications were addressed, and strategies to implement the changes were discussed based on the participant’s individual circumstances. During the first session, participants watched a cooking demonstration video in which a chef prepared a vegan meal. Strategies to enhance the flavor and appeal of plant-based meals were presented. Participants in TR had an unrestricted K^+^ diet.

### Physical fitness methods

Based upon the results of the FMS, individuals were advised as to the types of exercises they should perform. The recommended exercises were demonstrated for each participant along with a set of stretches. Participants in TR received a packet with instructions and illustrations of how to correctly perform the recommended movements. They were also instructed to walk for at least 30 minutes, 5 times per week at a moderate intensity to comply with standard physical activity recommendations(9). Participants were instructed to avoid prolonged periods in sedentary behavior (SB) by aiming to reduce sitting time (13).

The study coordinator attempted to contact the participants weekly during the intervention period to review the exercise program and to discuss any challenges that they might have encountered.(14). Participants were encouraged to contact the study coordinator if they had questions or needed clarification regarding their home-based exercise program.

### Pharmacy

Participants were specifically asked to bring all prescription (including those only taken as needed), and over-the-counter medications with them on test days (see Table 1). Participants provided their medications to the pharmacist who documented each medication including dosage directions and evaluated each medication for nephrotoxic effects, interactions, contraindications, and medication appropriateness in CKD. At BL, this was done for each patient without the pharmacist’s knowledge of patient group assignment. Any potentially nephrotoxic medication was addressed with the Medical Director of the study.

Once participants were randomized, those assigned to UC were excused without pharmacy associated changes as long as they were not taking medications identified as potentially harmful or nephrotoxic. Participants in TR met individually with the pharmacist. Based upon the information gathered, the pharmacist made evidence-based suggestions as part of the intervention. Medication schedule was based on pharmacokinetic and pharmacodynamic considerations as well as therapeutic guideline recommendations. Participants were educated about medication indication, benefit of nocturnal dosing of at least one antihypertension medication, and over-the-counter and herbal medications to eliminate or avoid (e.g., NSAIDs). PharmD investigators utilized study blood pressures and lab results to recommend initiation or dosing changes for ACEI, ARBs, and Aldosterone blocking agents. These recommendations were provided to the participant’s nephrologist for approval.

Participants indicated for patiromer, per study protocol, received education about the drug. The administration of the patiromer complied with the package insert. The pharmacists designed medication schedules incorporating patiromer and allowing for appropriate spacing from other medications.

For participants in TR, the pharmacist utilized the following algorithm based on the 2012 KDIGO guidelines (15) and with medical provider approval:

1. If the participant was on a RAS blocker, the dose was titrated up to a maximal dose in order to achieve a BP <130/80.
2. If the patient had an elevated urinary albumin excretion, the RAS inhibitor dose was also titrated to the maximal dose.
3. If K^+^ was >5.2 mEq/L, patiromer was provided and dosed according to the package insert.

### Behavior

From a behavior change perspective, the key features of the intervention were: an assessment of current physical activity and dietary habits, an assessment of the perceived benefits, barriers, and costs of behavior change, and patient participation in goal setting and developing personalized strategies to overcome barriers and prevent relapse. Regular self-assessment was used to empower participants to take responsibility for their own decisions and to make informed choices.

The study coordinator aimed to make follow-up contact with the participants on a weekly basis post-assessment, however she was unable to reach some participants as scheduled. This contact focused on the extent to which participants had achieved their goals since last contact, and offered reinforcement and assistance in problem-solving utilizing additional strategies as appropriate. Participants also received written materials tailored to their current concerns.

### Statistics

Data on the primary outcomes at 6 months (M6) were analyzed using a 2 (group: TR/UC) x 2 (time: baseline/M6) mixed factor ANOVA, with a focus on the interaction between group and time. Similar separate analyses were conducted at secondary endpoints of M1 and M3. All data were screened for violations of assumptions for these statistical tests. The alpha level was set at p < 0.05 for all analyses.

## Results

### Patient characteristics

Forty-two patients consented and started the study (see Figure 1 consort diagram). The baseline characteristics and demographic data of the participants are presented in Table 2. Following randomization, 27 were assigned to TR and 15 to UC. Analyses were conducted on 37 participants since 5 were eliminated due to only having BL data. Over the course of the study there were no changes in any anthropometric variables (see Table 3).

**Figure 1.** CONSORT DIAGRAM depicting the recruitment of participants for the study.

### Physical function

The intervention had no effect on any of the functional measures or indices of muscle strength or power (see Table 3).

### Sedentary time

The assessment of sedentary time was completed at BL, M3 and M6. Participants in both group
s spent 67-69% of their waking day being sedentary. This did not change over time and was not different between the two groups (See Table 3)

### Cardiovascular Function

There was a reduction in both brachial (bSBP) and central systolic blood pressures (cSBP) at M3 (p = .01) in TR, which was not apparent at M6. Central diastolic blood pressure (cDBP) was reduced by M3 while brachial diastolic blood pressure (bDBP) was not different between the two groups at M3. M6 readings were not different from BL or between groups. Central pulse pressure values did not differ over time or between groups during the duration of the study. Augmentation index (AIx@75) did not change from BL to M6 in TR but dropped in UC (p < 0.05) (see Table 3).

### Nutritional information

Total caloric intake (kcals) was reduced in TR at M6 (p = 0.03), but not in the UC group. Protein intake did not differ over time or between groups during the study. Sodium (mg) intake was different between groups at M1 with the UG group increasing their intake while TR had a modest reduction. Phosphorus (mg) intake trended downwards at M6 compared to BL in TR compared to the UC group (p=0.05) (See Table 3).

### Blood parameters

Serum K^+^ (mmol/L) remained stable throughout the study in both groups. Though not reaching statistical significance, serum Phos (mg/dl) levels trended downward in TR by M6, while there was a tendency for it to rise in UC. Neither IL-6 nor CRP changed over the study (see Table 3).

### Behavioral information

Neither self-efficacy to adhere to exercise nor self-efficacy to adhere to dietary recommendations changed over time or between groups over the course of the study. Of the five subscales of the KDQOL, the Effects of Kidney Disease on Daily Life showed significant findings (p=0.02). Changes in Effects of Kidney Disease on daily life for TR compared to UC did not show a significant difference in change from BL to M1 but did show a significant difference for the change from BL to M3 and M6. UC decreased from BL to M3 and M6, while TR increased. (See Table 3).

### Medication use

Eleven participants in TR were administered patiromer, according to administration guidelines, and 2 stopped taking their ACE/ARB medication at 3 months. Of the 37 participants with data in the study, 5 had changes to their ACE/ARB medications across the 6-month time period. One participant in UC stopped taking their ACE/ARB medication between 3 and 6 months. Two participants who were in TR and were not taking patiromer, at any time during the study started taking ACE/ARB at the 3-month assessment and continued through to M6.

## Discussion/Conclusion

The current study was primarily designed to determine the effect of an individualized, comprehensive, lifestyle intervention program including dietary, physical activity, pharmaceutical, and behavioral counseling on physical function, cardiovascular function, diet, and HRQOL in a sample of pre-dialysis CKD patients. We found that within the timeframe of this study, our intervention had no measurable effect on physical function, modest effects on cardiovascular function and nutritional intake but a major impact upon HRQOL in our sample of patients with CKD.

The most notable finding in this study was the significant and sustained impact of the intervention on self-reported effects of CKD. Efforts to make health care more patient-centered require greater knowledge of patient-reported outcomes, such as quality of life (QOL), where information comes directly from a patient without clinician interpretation. Such information provides meaningful insight as to the impact of disease and therapeutic interventions on daily life. CKD patients place tremendous value on their HRQOL, even over survival, and reportedly use impact on QOL to guide their therapeutic choices (16)

The QOL of CKD patients is a frequently overlooked yet critical consideration when evaluating their overall medical care (17). People living with CKD experience a high symptom burden with progressively impaired physical function and physical activity levels which can negatively affect HRQOL(18). In this study we found that CKD patients randomized to TR experienced significant improvements in their ability to live with CKD in their daily life. Positive changes that are clinically meaningful were also found for other aspects of HRQOL, including mental functioning, symptoms and problems associated with CKD and the perceived burden of CKD. Interestingly, these improvements in HRQOL were reported even in the absence of significant changes in physical function.

Most lifestyle studies involving CKD participants have focused on supervised exercise in association with dietary modification(3)(6). Howden et al. (5) utilized a comprehensive approach, involving frequent patient contact. They used a combination of supervised training (8 weeks) followed by a 10-month home-based program. This hybrid program produced improvement in the 6MWT distances in the intervention group. In contrast, our 6-month home-based program resulted in no improvements in any functional measures. However, despite this, we still observed meaningful changes in HRQOL.

Another notable finding in this study was the reduction in bSBP and cSBP that occurred at M3 in TR. Such a reduction in blood pressure is medically important due to the role that elevated blood pressure plays in the development of cardiovascular disease and progression of CKD. However, this reduction in blood pressure was not statistically significant at M6. We speculate that the gap in direct patient contact between M3 and M6 may have had an impact upon the attenuation of the BP reduction that was observed in TR by M6.

The pattern of change in blood pressure that was observed in this study is similar to what has been reported by Meuleman et al. (19) regarding Na restriction in a group of CKD participants. Similar to our study, Meuleman et al. (19) used a behavior change approach to get CKD participants to reduce their Na intake. As was seen in this study, the results of their intervention diminished between M3 to M6. In the current study, participants were contacted by the study coordinator over the phone or via email during the M3 to M6 period but there was no other direct in-person contact. Perhaps our findings suggest that there is need for study personnel to have direct contact with the participants to increase the probability of the latter adhering to the positive behavioral changes made. In contrast to the poor adherence to the home-based programming that we and others report, Van Craenenbroeck et al. (20) (21)reported excellent compliance to their 12-week, home-based exercise program. This could have been influenced by the fact that participants in the intervention group received an exercise bike (21).

One unusual finding in this study was a decrease in AIx@75 in UC at M6 with no change in TR. AIx@75 is a surrogate measure of arterial stiffness(3)which is a positive non-traditional risk factor for CVD (21). Researchers have reported mixed findings regarding the impact of exercise training on indices of arterial stiffness. In supervised studies lasting from 16 weeks to 12 months, researchers have found little evidence for exercise training affecting arterial stiffness as measured by either pulse wave velocity, or AIx75(3) (21). However, Mustata et al.(22)have reported a reduction in AIx in a sample of CKD participants after a 12-month hybrid supervised/home based program.

The intervention resulted in some important changes in nutritional intake. Participants randomized to TR decreased their intake of Na and by M6 they also consumed 300 mg less Phos than at BL. This change in Phos intake was reflected in their serum Phos levels which trended downwards at M6. Participants had been counseled to decrease their intake of animal protein but from the analyses performed it was impossible to decipher their protein source, (i.e., animal or plant). No restriction was placed upon their K^+^ intake which we assumed would be higher if they transitioned to a more plant-based diet.

Another observation that was noted in those randomized to TR was evidenced by the fact that at M6 participants in TR consumed 346 kcals less than at BL. The change in kcals, however was not enough to translate into changes in body weight or body composition. This is in contrast to what Howden et al. (5)found for their intervention at the 6-month mark.

Over the course of the study we carefully monitored both the doses and types of medications used by the participants. We found a slight increase in the use of ACE/ARB in TR with a slight decrease in UC. These changes did not reach statistical significance but the observation of a trend toward higher use of ACE/ARBs in TR, might suggest that the availability of patiromer to this group had an influence.

A number of limitations should be considered when interpreting the results of this study. The study duration was relatively short; a longer time period is needed to see changes in some of the parameters examined. We opted to use a totally home-based approach, thereby attempting to assess the effectiveness of our approach under ‘real-world’ (unsupervised) conditions. However, this meant that our results were affected by patient compliance to the tailored recommendations and though the study coordinator made a great effort, it was difficult to contact all patients as frequently as we had intended(8).

In conclusion, we have shown that a comprehensive, individualized, multicomponent program with behavioral counseling and close patient follow-up enables patients with CKD to feel better about managing their disease even without substantial changes in physical function. We believe this finding is unique; previously reported improvements in HRQOL following lifestyle intervention programs are coupled with changes in physical function (23). We also report modest changes in blood pressure and some aspects of nutritional intake, but patients feeling less bothered by their disease was the most important finding of this short trial. The importance of HRQOL is underscored by the association between QOL measures and mortality and hospitalization rates in dialysis patients(24). Future long-term studies (at least 24 months) are needed to explore the possibility that a comprehensive lifestyle intervention might result in favorable changes to key physiological parameters that impact upon cardiovascular disease and the progression of CKD, in addition to sustained improvements in the HRQOL of CKD patients.

## Data Availability

The available data are presented in the paper

## Acknowledgements

The authors would like to thank the participants who completed this study. We would also like to acknowledge the contribution made by Dr. Richard Wood in the original conceptualization of the study. Joy Whitback at RTANE was also instrumental in assisting with managing this study. The following students also assisted with data collection; Sam Santich, Kyle Leyshon, Jill Roach, Anna Platz, Phillip Dumont, Suzanne Pottratz, and Nedu Izuegbunam

## Authors’ Contributions

**S H**, along with MG was the principal investigator for this study. He coordinated all aspects of the study and took the lead in writing up the study results

**J H** was responsible for coordinating all behavioral aspects of this study. She was also heavily involved in the interpretation of the data and in manuscript preparation.

**B T** was responsible for the administration of the functional movement screen and the creation of the home-based fitness programs for all participants in the treatment group.

**M O** was a Pharm D professional who reviewed all medications taken by participants to make sure they were not nephrotoxic. For those who needed Patiromer she also instructed them as to how to correctly take this medication. She assisted in providing recommendations for changes to the medication regimen to the medical provider in the study as well as assessed all participants for adherence.

**C. D-C** was a Pharm D professional who performed an identical role to MO. She reviewed all medications taken by participants to make sure they were not nephrotoxic. For those who needed Patiromer she also instructed them as to how to correctly take this medication. She assisted in providing recommendations for changes to the medication regimen to the medical provider in the study as well as assessed all participants for adherence.

**A C** assisted with the study design and with the randomization of participants. He was responsible for performing all of the statistical analyses performed in this study. He was also instrumental in assisting with manuscript preparation.

**K D** was one of the study coordinators whose primary responsibility was assisting with data collection. She worked to help train and coordinate the involvement of other individuals who assisted with testing.

**J S** was the study coordinator who was responsible for contacting the participants, scheduling their visits, communicating with them between visits and assisting with all aspects of data collection. She has also assisted with manuscript preparation.

**E M** assisted with the nutritional aspects of the study. She coordinated the diet analyses and along under the supervision of the RD assisted with providing diet education to the participants.

**E E** was the person primarily responsible for recruiting participants for this study. She was also involved with data collection and manuscript preparation.

**B W** was the registered dietitian who helped to educate the participants on adopting a plant-based diet. She worked with other members of the team to educate the participants on what was required to adhere to the dietary recommendations.

**C S** was responsible for coordinating the assays performed in this study. She also assisted with manuscript preparation.

**B W** assisted with the strength assessments using the Cybex dynamometer. He performed this task throughout the study.

**M G** along with S H was the principal investigator for this study. He took medical responsibility for this study. He was heavily involved in the design of the study and the interpretation of the results.

## References

1. United States Renal Data System. 2017 USRDS annual data report: Epidemiology of kidney disease in the United States. Bethesda Maryland; 2017.

2. Navaneethan SD, Fealy CE, Scelsi AC, Arrigain S, Malin SK, Kirwan JP. A Trial of Lifestyle Modification on Cardiopulmonary, Inflammatory, and Metabolic Effects among Obese with Chronic Kidney Disease. Am J Nephrol [Internet]. 2015;274–81. Available from: http://www.karger.com/?doi=10.1159/000441155

3. Howden EJ, Leano R, Petchey W, Coombes JS, Isbel NM, Marwick TH. Effects of Exercise and Lifestyle Intervention on Cardiovascular Function in CKD. Clin J Am Soc Nephrol [Internet]. 2013 Sep [cited 2013 Sep 13];8(9):1494–501. Available from: http://www.ncbi.nlm.nih.gov/pubmed/23970136

4. Dutton GR, Lewis CE. ScienceDirect The Look AHEAD Trial□: Implications for Lifestyle Intervention in Type 2 Diabetes Mellitus. 2015;8:4–10.

5. Howden EJ, Coombes JS, Strand H, Douglas B, Campbell KL, Isbel NM. Exercise Training in CKD: Efficacy, Adherence, and Safety. Am J Kidney Dis [Internet]. 2015;65(4):583–91. Available from: http://linkinghub.elsevier.com/retrieve/pii/S0272638614012803

6. Ikizler TA, Robinson-cohen C, Ellis C, Headley S, Tuttle K, Wood R, et al. Metabolic Effects of Diet and Exercise in Patients with Moderate to Severe CKD□: A Randomized Clinical Trial. 2018;1–11.

7. Chen P, Huang Y, Kao Y, Chen J. Effects of an Exercise Program on Blood Biochemical Values and Exercise Stage of Chronic Kidney Disease Patients. J Nurs Res. 2010;18(2):98–106.

8. Hiraki K, Shibagaki Y, Izawa KP, Hotta C, Wakamiya A, Sakurada T. Effects of home-based exercise on pre-dialysis chronic kidney disease patients□: a randomized pilot and feasibility trial. 2017;1–8.

9. Reibe, D, Ehrman, J, Liguori G, & Magal M. ACSM’s Guidelines for Exercise Testing and Prescription. 10th ed. Deborah Riebe, Jonathan Ehrman, Gary Liguori MM, editor. Philadelphia: Wolters Kluwer; 2018.

10. Hays RD, Kallich JD, Mapes DL, Coons SJ, Carter WB. Development of the kidney disease quality of life (KDQOL) instrument. Qual Life Res. 1994;3(5):329–38.

11. Bandura A. Self-efficacy: the exercise of control [Internet]. Vol. 35, Choice Reviews Online. New York: Freeman; 1997. 35-1826–35-1826 p. Available from: http://www.cro3.org/cgi/doi/10.5860/CHOICE.35-1826

12. Bandura A. Guide for constructing self-efficacy scales. In: (Eds.)., Pajares F& UT, editor. Self-efficacy beliefs of adolescents,. Greenwich: Information Age Publishing; 2006.

13. Stephens SK, Winkler EAH, Trost SG, Dunstan DW, Eakin EG, Chastin SFM, et al. Intervening to reduce workplace sitting time□: how and when do changes to sitting time occur□? 2014;7:1037–42.

14. Karapolat H, Demir E, Bozkaya YT, Eyigor S, Nalbantgil S, Durmaz B, et al. Comparison of hospital-based versus home-based exercise training in patients with heart failure: effects on functional capacity, quality of life, psychological symptoms, and hemodynamic parameters. Clin Res Cardiol [Internet]. 2009;98(10):635–42. Available from: http://www.ncbi.nlm.nih.gov/pubmed/19641843

15. Kidney Disease: Improving Global Outcomes (KDIGO) CKD Work Group. KDIGO 2012 Clinical Practice Guideline for the Evaluation and Management of Chronic Kidney Disease. Kidney Int Suppl [Internet]. 2013;3(1):4–4. Available from: http://www.kdigo.org/clinical_practice_guidelines/pdf/CKD/KDIGOCKD-MBDGLKISuppl113.pdf%5Cn http://www.nature.com/doifinder/10.1038/kisup.2012.73%5Cn http://www.nature.com/doifinder/10.1038/kisup.2012.76

16. Garg AX, Suri RS, Eggers P, Finkelstein FO, Greene T, Kimmel PL, et al. Patients receiving frequent hemodialysis have better health-related quality of life compared to patients receiving conventional hemodialysis. Kidney Int [Internet]. 2017;91(3):746–54. Available from: http://dx.doi.org/10.1016/j.kint.2016.10.033

17. Kimmel PL, Cohen SD, Weisbord SD. Quality of life in patients with end-stage renal disease treated with hemodialysis□: survival is not enough□! 2008;21(suppl 13).

18. Mackinnon HJ, Wilkinson TJ, Clarke AL, Gould DW, Sullivan TFO, Xenophontos S, et al. The association of physical function and physical activity with all-cause mortality and adverse clinical outcomes in nondialysis chronic kidney disease□: a systematic review. 2018;

19. Meuleman Y, Hoekstra T, Dekker FW, Navis G, Vogt L, Boog PJM Van Der, et al. Sodium Restriction in Patients With CKD: A Randomized Controlled Trial of Self-management Support. 2018;69(5):576–86.

20. Van Craenenbroeck a. H, Van Craenenbroeck EM, Kouidi E, Vrints CJ, Couttenye MM, Conraads VM. Vascular Effects of Exercise Training in CKD: Current Evidence and Pathophysiological Mechanisms. Clin J Am Soc Nephrol [Internet]. 2014;9(7):1305–18. Available from: http://cjasn.asnjournals.org/cgi/doi/10.2215/CJN.13031213

21. Van Craenenbroeck AH, Van Craenenbroeck EM, Van Ackeren K, Vrints CJ, Conraads VM, Verpooten GA, et al. Effect of Moderate Aerobic Exercise Training on Endothelial Function and Arterial Stiffness in CKD Stages 3-4: A Randomized Controlled Trial. Am J Kidney Dis [Internet]. 2015;66(2):1–12. Available from: http://linkinghub.elsevier.com/retrieve/pii/S0272638615005946

22. Mustata S, Chan C, Lai V, Miller JA. Impact of an exercise program on arterial stiffness and insulin resistance in hemodialysis patients. J Am Soc Nephrol. 2004 Oct;15(10):2713–8.

23. Aioke DT F B M K, Ammirati, A CL. Home-based versus center-based aerobic exercise on cardiopulmonary performance, physical function, quality of life and quality of sleep of overweight patients with chronic kidney disease. Clin Exp Nephrol. 2018;22:87–98.

24. Mapes DL, Lopes A, Satayathum S M, Goodkin D, Locatelli F, Fukuhara S YE, Kurokawa K, Saito A, Ä Bommer R WR, Held H and PF. Health-related quality of life as a predictor of mortality and hospitalization: The Dialysis Outcomes and Practice Patterns Study (DOPPS). Kidney Int. 2003;64:339–49.

